# Short time effect of Covid 19 pandemic on HbA1c and acute metabolic complications in children with type 1 diabetes

**DOI:** 10.1101/2021.04.18.21255687

**Authors:** Ilknur Arslanoglu, Figen Akcali, Fatma Yavuzyilmaz, Mehmet Ali Sungur

## Abstract

**Background:** COVID19 pandemic is currently affecting every aspect of daily life of communitiesy throughout the world. We aimed to check how this situation affects the metabolic control of children with type 1 diabetes.

**Methods:** We analyzed all patients with type 1 diabetes a HbA1c test after at least two months ensuing the start of the epidemic in Turkey. We compared the results with the most recent HbA1c test in the hospital’s automation system before the epidemic. In addition, diabetic ketoacidosis (DKA) and severe hypoglycemia rates were compared.

**Results:** Among the eligible 219 cases 77.6% had decreased HbA1c levels according to their former result. Mean drop was about 9.71% compared to the former test in the whole group. Age, sex and time interval between two tests were not found to affect this tendency. Diabetic ketoacidosis rate was the same as before the pandemic, whereas severe hypoglycemia rates increased.

**Conclusions:** Despite the potential of the pandemic to affect routine care of chronic diseases in a negative way the short term metabolic control of type 1 children with type 1 diabetes improved. Telemedicine support by the diabetes team and increased care in the family environment might be possible explanations.

**Grafik Abstract:** 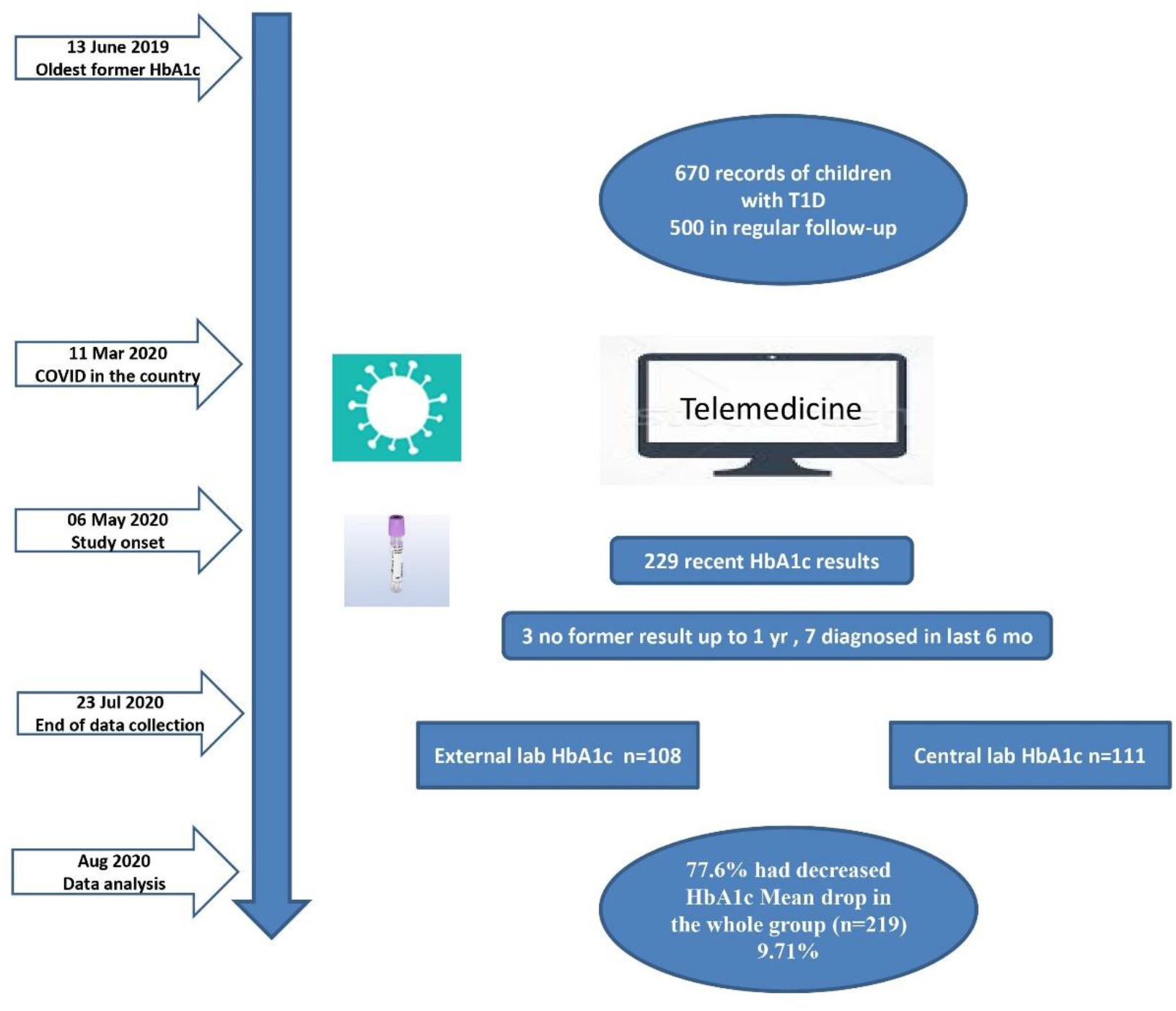

**What is already known on this topic?:** Care of chronic diseases is a concern in the recent COVID-19 pandemic era.

**What this study adds?:** A marked improvement in HbA1 C levels of children with Type 1 diabetes in short term during COVID-19 pandemic was noted suggesting factors including closer follow-up by diabetes care team using telemedicine and more family support might be responsible.

## Introduction

COVID19 pandemic is affecting every aspect of daily life throughout the world. The healthcare system is expectedly at the center of the multidirectional struggle against the virus and still responsible for the remaining health problems (1). With the pandemic huge organizational changes happened regarding the care of chronic diseases although differences exist between countries (2). Some of the changes are spontaneous and uncontrollable (neglecting routine care and control visits due to panic and lack of facilities), some on the other hand are conscious and systematic (postponing some elective health care services by the government, increasing importance of telemedicine). Education in the medicine targeting professionals, students, patients or the general public at large has shifted to online facilities as well (3). Countless webinars were conducted in which the main topic initially was the viral disease itself and several other medical issues thereafter.

In our pediatric diabetes center too we changed our type 1 diabetes follow-up strategy after the first COVID case was reported on March 11^th^ in Turkey. At the beginning we expected serious burdens of the epidemic on our patient’s health. These were the infection risk itself on one hand and the deterioration of metabolic control, increase in blood sugars and occurrence of acute complications like severe hypoglycemia and DKA on the other hand (4). In time we started collecting glucometer reports, diary records, locally measured HbA1c results and finally started accepting clinic visits in a limited fashion. Our impression was that the metabolic control of the majority did not worsen, even improved, prompted us to design the present study. In this study we compared the HbA1c of individuals and the DKA and severe hypoglycemia rate of the whole cohort before 11 March 2020 and after 01 May 2020.

## Participants and Methods

### General setting of patient care before and during epidemic

Our center is part of the Pediatric Endocrinology Department in a university hospital. It is one of the registered SWEET centers as well (5). Before the epidemic we checked patients with type 1 diabetes in every three months in average in the outpatient clinic and during a complete visit day the family met diabetes nurse, dietician, social worker, pediatrician, pediatric endocrinologist separately and attended additionally the motivational group interviewing going on since ten years uninterruptedly. The background population comprises of 670 recorded patients with type 1 diabetes and approximately 500 of them are still on follow-up with >50% on insulin pump therapy.

During the epidemic a pandemic ward was established in our hospital immediately and all non-emergent services stopped. We stopped all the above mentioned activities as well. Instead we encouraged families to send diaries, downloads of glucometer, CGM and pumps. As our diabetes team had formerly tight communication with families of children with type 1 diabetes via social media and personal e-mail &phone we made group announcements suggesting not to come to the hospital, rather communicate distantly but in an increased frequency. Formerly we had one Facebook account and five Whats App (WA) groups. We established three additional WA groups, two for old and new pump users and one for recently diagnosed patients. We asked the patients to share every hypoglycemia event whether severe or not and their intervention. In addition regular diary checks were made and every other event needing consultation related to diabetes or not was shared in the groups. But private communication of the patients with all team members is accepted limitless if the patient prefers to do so. Using this way we adjusted insulin dosages, pump settings and reeducated them especially for carbohydrate counting and advanced insulin management. Moreover we prepared videos for education, for special day celebrations, etc. In addition we organized serial zoom meetings for pump users.

This is a retrospective analysis of children with T1DM who had a HbA1C level drawn between May 6th to July 23rd 2020. The data were analyzed retrospectively for the change in HbA1C levels and the frequency of reported DKA or hypoglycemic episodes. Only 219 children with T1DM were eligible to be included in this study due to lack of available HbA1C data prior to the COVID 19 pandemics. The study was conducted in concordance with the Declaration of Helsinki-Ethical Principles for Medical Research Involving Human Subjects. The study was approved by the Ethics Review Board (protocol number: 2020/195). Since our study was retrospective, no informed consent was taken.

### Inclusion criteria

Having a diagnosis of type 1 diabetes before the age of 18, being checked for HbA1c between the dates May 1^st^ and July 23 2020 (last day of data collection).

### Exclusion criteria

Absence of a former HbA1c test up to one year before and have been diagnosed in the last 6 months (to exclude the hba1c level before insulin therapy).

### Data collection

#### Demographics

Data for age and sex were obtained from hospital files.

#### HbA1c

We evaluated every subject with type 1 diabetes who has a HbA1c check after the date of May the first 2020. Some of the tests were performed in our hospital, some of them locally and the report was sent to us via internet. We created the list for all these patients and then checked the hospitals automation system for their most recent HbA1c test before 11 March 2020. We recruited patients with a HbA1c measurement up to one year back according to the post-pandemic test. We analyzed the change in HbA1c before and after the epidemic in the total group and in subgroups. Our test results are reported in DCCT units. We classified patients as improved, persisted and deteriorated with respect to HbA1c, persisted was defined as exactly the same result before and after compared up to one decimal digit.

#### Diabetic ketoacidosis and severe hypoglycemia

We routinely collect data for acute complications for SWEET entries. Diabetic ketoacidosis is defined as a diabetic ketosis with a measured blood pH under 7.30 and severe hypoglycemia as the status needing another person’s assistance to force sugar intake or glucagon injection or intravenous glucose (6,7). Diabetic ketoacidosis was analyzed after exclusion of new onset cases, whereas severe hypoglycemia comprised of all cases provided that the case is included in the SWEET registry in order to make a comparison with the former year possible. We compared last one year’s (January 1^st^ 2019-December 31^st^ 2019) rates with those of March 11^th^-July 11^th^ 2020 multiplied by three.

## Statistical Analysis

Change in HbA1c was expressed as the percentage of the former result. Descriptive statistics were given as mean ±standard deviation for numerical variables and as frequency and percentage for categorical variables. Kolmogorov-Smirnov test was used to examine normality assumption of numerical variables. Paired samples t-test was used to analyze before and after values. Independent samples t-test and Mann-Whitney U test or One-Way ANOVA and Kruskal-Wallis test were used to compare groups based on the group numbers compared and normality assumption. Categorical data were analyzed with Pearson chi-square or Fisher-Freeman-Halton test, as appropriate. Data were analyzed with SPSS v.22 statistical package, and a value of 0.05 was considered as statistical significance level.

## Results

First of all, we did not face a suspected or proven COVID-19 infection among our patients with type 1 diabetes.

We collected 229 HbA1c results between May 6 and July 23, 2020. Three patients had no former test up to one year back and 7 patients were diagnosed during last 6 months, these subjects were excluded from the analysis. There remained 219 eligible subjects. Number of the patients who visited our hospital after May 1^st^ is 111 (50.7%). The number of those who sent us their HbA1c report performed locally after the same date is 108 (49.3%). Table 1 shows general characteristics of the studied subjects.

**Table 1.** General characteristics of the subjects having tested for HbA1c before and after the pandemic

|  | Overall (n=219) | Local* HbA1c test (n=111) | External** HbA1c test (n=108) |
| --- | --- | --- | --- |
| Sex, n (%) |  |  |  |
| Male | 101 (46.1) | 61 (55.0) | 40 (37.0) |
| Female | 118 (53.9) | 50 (45.0) | 68 (63.0) |
| Age, mean±SD (min-max) | 12.59±4.31 (1-23) | 13.10±4.52 (2-23) | 12.06±4.03 (1-23) |

\*Test after pandemic was performed in our hospital
\*\* Test after pandemic was performed in another hospital where the patient is resident and report was sent via internet

Table 2 shows HbA1c results before and after. HbA1c after the epidemic was statistically significantly lower in the whole group (p<0.001). The mean drop amount was 9.71% of the first results. When analyzed according to the site of testing, there were statistically significant drops both for local and external laboratory subgroups (both p values were <0.001). The mean drop amount was 9.97% of the first results for local group and 9.34% for external group (Table 2).

**Table 2.** HbA1c results before and after the exposure to the pandemic

| HbA1c (%) | Overall<br>(n=219) | Local HbA1c test (n=111) | External HbA1c test (n=108) |
| --- | --- | --- | --- |
| Before | 8.55±1.57 | 8.63±1.69 | 8.46±1.44 |
| After | 7.72±1.35 | 7.77±1.42 | 7.67±1.27 |
| p | **<0.001** | **<0.001** | **<0.001** |

The time interval between two tests was 72 - 373 (median 140) days.

We analyzed further subgroups according to time interval (<90 days, 90-120 days, 121-180 days, >180 days). There were 10 (4.6%) patients with an interval <90 days between two results, while there were 55 (25.1%) for 90-120 days group, 107 (48.9%) for 121-180 days group, and 47 (21.5%) for >180 days group. The changes for HbA1c results in these four groups were given in Table 3. There were statistically significant differences between two results for 90-120 days, 121-180 days, >180 days interval groups (all p values were <0.001), except <90 days interval group. Although there was a decrease of 4.8 % for this group in the same way, it was not found statistically significant probably due to the limited number of subjects (p=0.307).

**Table 3.** HbA1c results before and after pandemic according to the time interval between two tests

| HbA1c (%) | <90 days<br>(n=10) | 90-120 days<br>(n=55) | 121-180 days<br>(n=107) | >180 days<br>(n=47) |
| --- | --- | --- | --- | --- |
| Before | 8.54±1.66 | 8.87±1.78 | 8.26±1.39 | 8.83±1.61 |
| After | 8.13±1.59 | 7.82±1.44 | 7.44±1.09 | 8.13±1.58 |
| p | 0.307 | **<0.001** | **<0.001** | **<0.001** |

We classified patients as improved, persisted and deteriorated and following rates were found respectively. There were 170 (77.6%) patients whose HbA1c levels were decreased according to their first result. The mean decrease was 1.23 (range, 0.1 to 4.6) in this group. There were 43 (19.6%) patients having increased HbA1c values with a mean increase of 0.62 (range, 0.1 to 4.1), and 6 (2.7%) patients whose HbA1c values were similar. Comparison of these groups in terms of age and sex was shown in Table 4. There was no significant difference between these three groups in terms of sex (p=0.705), age (p=0.709) and the time interval between two results (p=0.618). Since there was a few patients in persisted group to compare, and can see the difference between the two groups better, when we compare only improved and deteriorated groups with each other, the similar results were obtained. Again there was no significant difference for sex (p=0.497), age (p=0.863) and the time interval between two results (p=0.450) between two groups.

**Table 4.** Comparison of improved, persisted and deteriorated patient groups with respect to the HbA1c results before and after the pandemic by means of demographics and time interval

|  | Improved<br>(n=170) | Persisted (n=6) | Deteriorated<br>(n=43) | p |
| --- | --- | --- | --- | --- |
| Sex, n (%) |  |  |  |  |
| Male | 81 (47.6) | 2 (33.3) | 18 (41.9) | 0.705 |
| Female | 89 (52.4) | 4 (66.7) | 25 (58.1) |  |
| Age, mean±SD (min-max) | 12.52±4.31 (1-23) | 14.00±3.41 (10-18) | 12.65±4.41 (4-23) | 0.709 |
| Time Interval, median (IQR) [min-max] | 135 (56) [72-373] | 149 (24) [116-160] | 147 (50) [81-340] | 0.618 |
| Time Interval, n (%) |  |  |  |  |
| <90 days | 6 (3.5) | 0 (0.0) | 4 (9.3) | 0.224 |
| 90-120 days | 47 (27.6) | 1 (16.7) | 7 (16.3) |  |
| 121-180 days | 79 (46.5) | 5 (83.3) | 23 (53.5) |  |
| >180 days | 38 (22.4) | 0 (0.0) | 9 (20.9) |  |

According to our center’s benchmarking report created by SWEET, DKA was observed in 3.6 cases/100 patient years and severe hypoglycemia in 0.2 cases/100 patient years in the year of 2019. In the four months of epidemic we fixed 5 cases of DKA and 4 cases of severe hypoglycemia with corresponding incidence rates of 3.6 and 2.8 /100 patient years respectively. In two severe ketosis cases not checked for blood pH, hospital admission was prevented via telemedicine. Thus, DKA remained exactly the same whereas severe hypoglycemia increased more than 10 times.

## Discussion

Our country faced with the epidemic slightly later then its neighbors. Although dead rates were not so high, case numbers and the impact on social life were quite high. Short after the first case schools and restaurants were closed and a few weeks later partial curfew was declared as well as travel between cities was forbidden.

The impact of the epidemic on the healthcare organization was big as we mentioned in the introduction section. There was a shift from elective and routine care to emergency care. There are reports of pediatric diabetes cases diagnosed for Covid-19 infection, but none of our patients faced the disease (8).

The reasons to have concerns about the management of children with type 1 diabetes were numerous. First of all in early days of the epidemic all chronic diseases were announced as high risk reasons by scientific authorities. In addition due to the interruption of routine care the patients who drop out from the follow-up could face metabolic difficulties. Moreover psychologic problems like depression and anxiety could occur (9). Since the curfew comprised of children physical activity could be limited. An appreciable measure taken by the government was that medications for chronic disease were affordable without necessary refreshments of official documents or receipts. Therefore we hypothesized that blood sugars of our patients and cases of DKA will increase.

By the time we observed the reverse picture and as the present study shows the short term metabolic control improved regarding the HbA1c levels. The mean drop is quite striking with almost a 10 % difference in comparison to the former value. On the other not all subjects have a falling HbA1c level, a relative small part of the whole group (19.6%) have increased levels. We could not explain this different response to exposure to the situation of epidemic by age, sex or time lapse between to tests. Now we are planning to analyze improving and deteriorating groups in more detail. For example diabetes duration, sudden changes in home environment or a steady worsening even before the epidemic might be possible explanations. Since we excluded new onset and honeymoon periods, there is no confounding reason for the improvement of the subjects in the corresponding group.

While we are surprised with our observation, looking at the very recent literature, we found similar observations in various studies with different samples of design and target populations (10–12). These studies are conducted in adult populations and reported changes are quite minor changes. On the other hand some other studies revealed worsening of glycemic control due to lockdown (13). There is a small study in patients with type 1 dibates age showing a slight improvement in “time in range” determined by CGM especially when physical activity was planned during pandemic (14).

Diabetic ketoacidosis rates remained the same. The comparison of a certain period of the year with the whole year might be problematic due to seasonal differences, but our analysis helps to obtain a crude estimation about the issue. Telemedicine again helped us to prevent more cases of DKA as similar observations in the literature (15). On the other hand severe hypoglycemia incidence rose strikingly with respect to former year’s data. This might be expected as a byproduct of improved sugar levels though this unwanted situation is less frequent using diabetes technology efficiently (16). Severe hypoglycemia incidence is still acceptable despite this relative striking rise. The incidence of severe hypoglycemic coma is currently reported as 3-7 per 100 patient years in various pediatric diabetes cohorts (17). In an adult study from Spain using standalone CGM, pre- and post-lockdown hypoglycemia rates were found similar, but they did not reported mean glycaemia or HbA1c during these periods (18).

Our diabetes team was fortunate that our announcements and suggestions were responded most positive than ever by our patients. Telemedicine was already one of our practices formerly. But during this period the majority of our team members worked flexibly and the time gained was spend for the care of diabetic patients. The increased online activity had a positive effect on our team’s patient care standards regarding technical issues and communication skills to our opinion. The advantage of group discussion via Whats App groups is quick reply since all the team is in the group, contribution of other patients in practical issues and supervision of the dialog by the senior team member.

We don’t think that the improvement is related solely to the team’s performance. More important than this, the family environment changed in a positive way, just the opposite what we expected. We can comment on this that how well organized the conditions in the external social life are, the families of children with type 1 diabetes and the child hardly can cope with it! For example there is a national “Diabetes in School” program conducted in our country since years with the contribution of the pediatric endocrinologists and of ministries of health and national education, so that the awareness in the schools is high about type 1 diabetes. But still, as some of our mothers of children with type 1 diabetes said: “During the epidemic I managed my child’s disease much better!”

Last but not least we can speculate about another effect of the epidemic: Such a collective treat might have some positive effects on people, sharing a common fear, watching officials and healthcare professional fighting for humans may awake feelings of responsibility. Going further, little ones with type 1 diabetes may think that there could be more important problems of life than diabetes.

## Study Limitations

Our interpretations for the reason of the positive change in HbA1c are depending on subjective opinions and some feed-back from the families. This might be a limitation of the study. Another limitation is the way of calculation of acute complications, where we compared real whole year data with calculated whole year data.

## Conclusion

The increased demand for emergency healthcare and psychosocial distressing factors might have negative effects on the routine care of children with type 1 diabetes. This potential seems to be overcome via remote support and increased time spent at home in the family environment. Studies designed using questionnaires might shed light on this issue.

## Data Availability

All data referred to in the manuscript is availabile.

